# Effectiveness and efficiency of pre-season administration of long-acting monoclonal antibodies for infants born to RSV vaccinated mothers: a modelling study

**DOI:** 10.64898/2026.06.16.26355774

**Authors:** Julia Mayer, Ayaka Monoi, Fabienne Krauer, Kevin van Zandvoort, Beate Kampmann, Matthieu Domenech de Cellès, Stefan Flasche

## Abstract

**Background:** A maternal vaccine (MV) and a long-lasting monoclonal antibody (la-mAB) have been licensed to protect children against RSV. Given the swift waning of their protection, we evaluated the added benefit of seasonal la-mAB administration for children born to vaccinated mothers shortly after the RSV season in Germany.

**Methods:** We fitted an age- and birth season-structured catalytic model to cross-sectional seroprevalence data of RSV antibodies using a Bayesian framework to estimate the timing of RSV infections in children <5. We then estimated the incidence of severe outcomes in the absence of immunisation in children <1. Finally, we estimated the impact of the MV and of additional la-mAB administration, accounting for the waning of protection.

**Results:** We estimate that children would, on average, be 7 months old (mo) at their first infection, those born in the autumn being the youngest at first infection (4 mo). Together with the children born in the winter, they account for 46% of all RSV hospitalisations and 62% of RSV ICU admissions in unimmunised <1 yo. MV would prevent a total of 776 (473-1,122) hospitalisations per 100,000 vaccinees and 73 (51-94) ICU admissions per 100,000 vaccinees, predominantly among autumn-born children. The summer birth cohort would benefit most from additional la-mAB administration, preventing an additional 46% (10-63) of ICU admissions compared to MV alone, corresponding to an additional 25 (4-45) ICU admissions prevented per 100,000 immunised children.

**Conclusion:** Compared to MV alone, the impact of MV+la-mAB on RSV hospitalisations and ICU admissions would likely be modest.

## Introduction

Respiratory syncytial virus (RSV) infection is the most common cause of severe respiratory tract infection in young children and the main cause of bronchiolitis in infants.^1^ While most RSV infections result in mild disease, about one-third of infants develop RSV-associated acute respiratory infections (RSV-ARI), leading to about 3 million hospitalisations and over 66,000 deaths worldwide in children under the age of five.^1,2^ It is estimated that most children will have experienced at least one RSV infection by the age of two years.^3^ In Germany, about 14,000 RSV-related hospitalisations were reported in this age group in 2023, mostly in children under the age of one.^4,5^

RSV epidemiology is typically very seasonal.^6^ In temperate climates like Germany, the virus starts circulating in colder months, between November and March, with the highest incidence observed in December to February, putting healthcare settings under pressure every winter.^7,8^ Children born during the RSV season or shortly before are at a higher risk of hospitalisation than those born in other months.^9,10^ Infection leads to short-term immunity in all age groups, including pregnant women. Anti-RSV antibodies are known to be transferred from pregnant mothers to their foetus across the placenta. Studies suggest that this grants some passive immunity against RSV disease in most children in the first months of life.^11,12^

Until recently, a short-acting monoclonal antibody (mAB), palivizumab, was the only immunisation option available against RSV. In Germany, it was recommended for high-risk infants and needed to be administered monthly in the RSV season. In 2022, a long-acting mAB (la-mAB), nirsevimab, was approved by the European Commission.^13^ Concurrently, a maternal RSV vaccine (MV), Abrysvo, was licensed in 2023.^14^ In June 2024, the Standing Committee on Vaccination (STIKO) recommended the use of the la-mAB, administered seasonally to infants, unless they are born to vaccinated mothers.^15^ At present, STIKO has not issued any recommendation on the use of the MV. Passive immunisation in infants via MV decays swiftly with potentially little protection left after six months.^16^ With a year-round approach, this implies that children born to vaccinated mothers at the tail-end of the RSV season, i.e. those who would otherwise not get the birth dose of la-mAB but instead be eligible for the catch-up in autumn, might be left without meaningful protection when they are exposed to RSV in their first full season. Administering the la-mAB to these children right before the start of their first RSV season could enhance their protection albeit at an age where their risk for severe disease is lower.

We estimated the hypothetical impact of offering la-mAB to German infants born outside the RSV season to RSV-vaccinated mothers. To do so, we developed a modelling framework to estimate the number of RSV-related hospitalisations and admissions to the intensive care unit (ICU) by season of birth in the absence of interventions, with maternal vaccination only and with maternal vaccination and added seasonal administration of la-mAB.

## Methods

### Data

#### Demographic data

We modelled a cohort of 692,989 children, in line with the size of the 2024 German birth cohort as published by the German statistical office.^17^

#### Seasonal RSV infection pressure

Given the absence of systematic RSV sero-surveillance data from Germany but similarities in RSV seasonalities, we estimated the age-specific proportion of RSV-seroconverted children less than one year old from the Dutch PIENTER-2 and PIENTER-3 studies.^18–20^ Briefly, these studies enrolled a total of 10,932 individuals of all ages from the Dutch population, and blood samples were collected for 10,236 of them. The blood samples were tested for anti-RSV IgG and IgA antibodies, and the titres were used to ascertain the participants’ seroconversion status: children younger than 500 days with IgA values above 0.2 AU/mL were classified as seroconverted, while a threshold of 1.0 AU/mL for IgG was used for older participants.^18^ Children under the age of 1 were oversampled and 817 had blood drawn. We divided the data into bi-monthly age groups and calculated the seroprevalence of past RSV infection, along with 95% binomial confidence intervals (CI), in each of the groups.

#### Age distribution of RSV disease burden

In the absence of accessible information on granular age-dependent RSV-associated hospitalisation incidence in German infants, we used the distribution of hospitalisations and ICU admissions in children <5 years derived as part of a literature review looking at the age distribution of RSV disease in low- and middle-income countries (LMICs).^21^

#### RSV disease burden

German hospitals are legally required to provide anonymised RSV case notification to the German Institute for the Hospital Remuneration System (InEK), but at a wider resolution than needed for our model.^4^ We calibrated the granular outputs of the age distribution above such that the total number of hospitalisations would match that reported in children <5 years with an RSV-specific primary diagnosis (ICD-10 codes J12.1, J20.5, and J21.0 which relate to RSV pneumonia, acute bronchitis due to RSV, and acute bronchiolitis due to RSV respectively)^22^ in 2023. We did not include secondary diagnoses as we were interested in hospitalisations primarily associated with RSV. Restricting the number of hospitalised cases to those admitted to the ICU gave us the number of ICU admissions in 2023.

#### Vaccine effectiveness

We used the procedure described by Monoi *et al.*, to estimate the effectiveness of the MV and of the la-mAB at a given time following immunisation.^23,24^ Briefly, monthly vaccine and immunisation efficacy estimates are taken from the respective phase 3 clinical trials^16,25^ and used to estimate the protection offered by the immunisation product at a given time point after administration. The waning rate of the efficacy is assumed to follow an Erlang-2 distribution and estimated using a Markov Chain Monte Carlo (MCMC).

### Model

We estimated the impact of RSV immunisation in three steps (Fig. 1). First, a catalytic model estimated the age- and season of birth-dependent cumulative incidence of RSV infection in the first year of life. Then we estimated progression rates to severe disease necessitating hospitalisation or ICU admission in Germany in the absence of immunisation. Finally, we estimated the impact of MV and subsequently of additional la-mAB administration.

**Figure 1:**
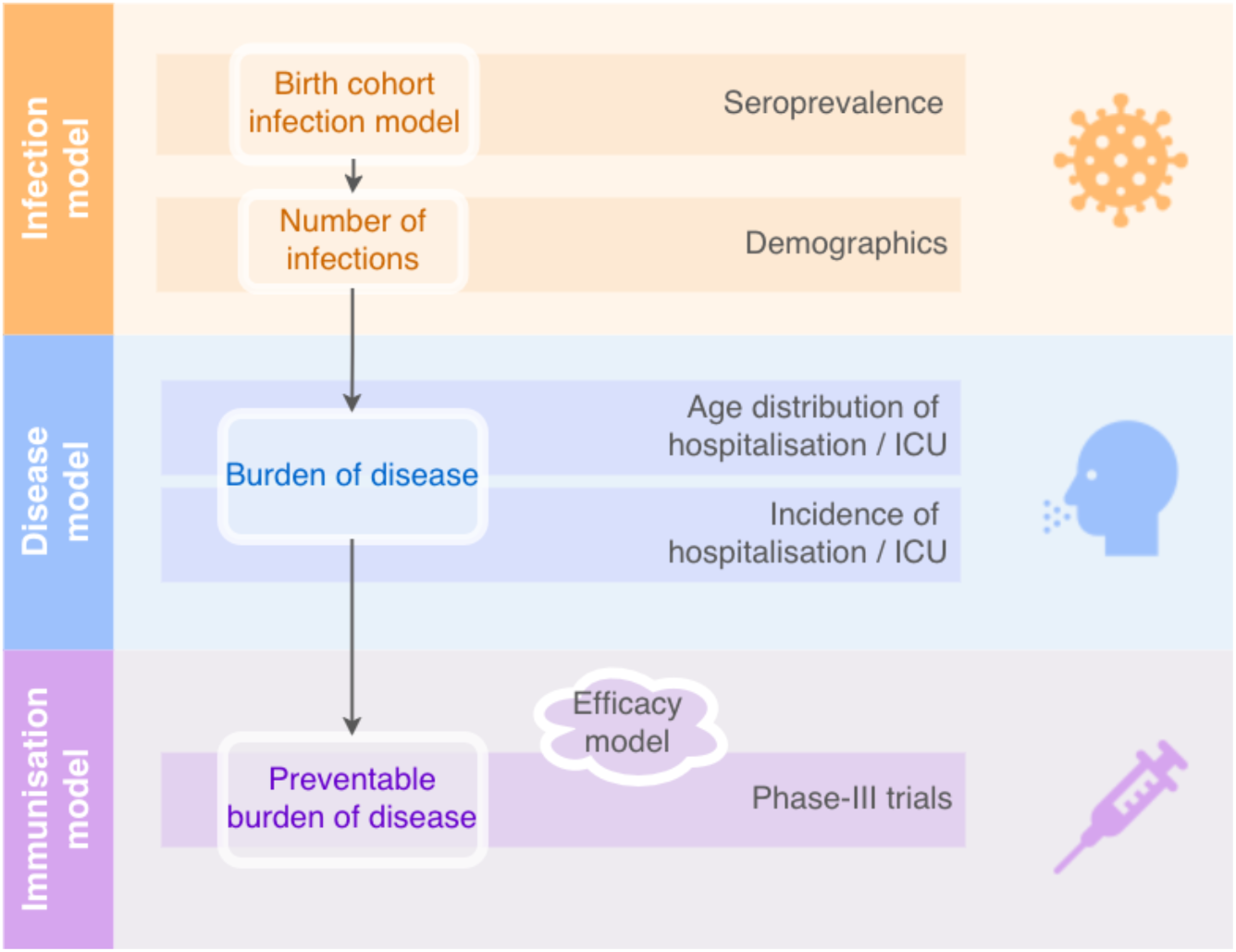
Model structure The model is divided into three parts: a catalytic infection model, a burden of disease model, and an immunisation model. ICU: intensive care unit admission

#### Infection model

We built a catalytic model to estimate the cumulative incidence of RSV-infection associated seroconversion at a given age and in dependence on the time of birth during the year. To account for the seasonality of RSV, children were divided into four groups based on their season of birth (summer, autumn, winter, spring). The seasons were defined as follows: summer lasted from June to August, autumn from September to November, winter from December to February, and spring from March to May. We also allowed the force of infection (FOI) to vary by season. The birth season then determined the age at which children were exposed to a specific seasonal FOI λ.

A proportion π of children was assumed to be born protected by maternal immunity which waned with a second order Erlang distributed rate ω. The resulting model had four compartments: M_1_ and M_2_ for the children protected by maternal immunity, S for those susceptible to infection (i.e., seronegative), and R for those who were infected, and no longer susceptible to RSV infection that season.

The model is governed by the following set of equations:

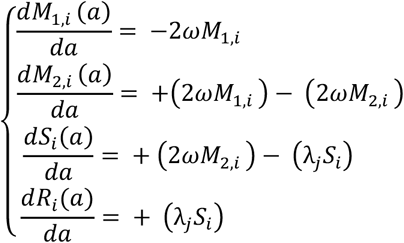

where a proportion π of newborns starts in M_1_ and the remainder in S. In the model, *a* indicates age, ω is the waning rate of maternal immunity, and λ_!_ is the FOI acting on a birth cohort *i* during season *j* that they reach at age *a*.

The proportion of children born with maternal immunity π, the waning rate of maternal immunity ω, and the four seasonal components of the force of infection λ_!_ were estimated by fitting the model to the Dutch seroprevalence data. Thus, we obtained the proportion of infected seroconverted children at a given age in the total population stratified by season of birth. Infection incidence estimates were then derived using Germany’s 2024 birth cohort size.

The model was built in R (version 4.4.1) using the odin2 package.^26^ The fitting was done using adaptative Metropolis Hastings sampling via MCMC from the monty package.^27^ We used a binomial log likelihood function for each of the four birth cohorts:

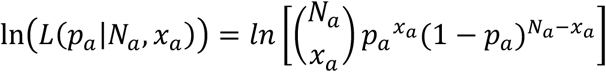

where *_pa_* is the proportion of seroconverted children at age *a* as estimated by the model, *_Na_* is the total number of children of age *a*, and *_xa_* is the observed number of seroconverted children at age *a* as given by the data. All parameters were estimated using Uniform prior distributions except for ω, for which we used a Normal distribution centred around six months based on the literature.^28–31^ More information on the fitting procedure, as well as parameter specifications, can be found in the appendix.

#### Burden of disease model

We used the posterior estimates of the age distribution for hospitalised, and ICU admitted RSV cases given by Mahmud *et al*^21^ to estimate the age distribution of RSV-hospitalisations and ICU admissions in German children < 5 years. We calculated progression rates from infection to severe disease by dividing the number of hospitalisations and ICU admissions by the age specific estimates for the number of RSV infections in the infection model. These disease progression rates were subsequently applied to the seasonally disaggregated estimates of infection incidence to estimate age- and season-specific disease burden.

#### Immunisation model

For each of the four seasonal birth cohorts, we derived the estimated number of cases averted by MV as the product of the vaccine effectiveness against hospitalisation granted by the MV at a given time since birth, i.e., at a given age, and the number of hospitalisations or ICU admissions at the same age that would have occurred in the absence of vaccine. We calculated the burden of disease that may be avertable with complete vaccine coverage. We assumed there would be no indirect effects from immunisation and only considered the effect on the infant, ignoring the benefit of MV for the mothers.

Similarly, we derived the estimated number of cases averted by adding nirsevimab administration as the product of the effectiveness of la-mAB at a time since administration with the residual disease burden in the presence of MV use. To model the seasonal catch-up, we assumed that the la-mAB would be given right before the start of the infant’s first RSV season: the winter-born cohort was immunised at 11 months of age, the spring-born cohort at 8 months, the summer-born cohort at 4 months, and the autumn-born cohort at 1 month.

We kept track of the number of doses administered and derived the number needed to immunise (NNI) to prevent one hospitalisation or ICU admission.

#### Uncertainty

To propagate the uncertainty in the parameters across the models we used bootstrapping. We ran 100 bootstraps, for each the results are based on (i) a single sample from the posterior distribution of the catalytic infection model, (ii) a single sample from the posterior of hospitalisations and ICU admissions age distribution, (iii) a single sample from the posterior of the MV protection trajectory, and (iv) a single sample from the posterior of the la-mAB protection trajectory.

We tested the sensitivity of our results to the assumption on the duration of naturally-derived maternal protection against infection and the shape of waning protection. Thus, we refitted the infection model using i) a non-informative prior, ii) a first order Erlang distribution (exponential), iii) a third order Erlang distribution, and iv) birth season-specific protection levels.

The combination of MV and la-mAB has not been tested in real life yet, so evidence on the type of protection generated by this approach is lacking. An ongoing trial recently reported similar AB titres among children born to unvaccinated mothers and those born to vaccinated mothers six weeks after administration of the la-mAB.^32^ Therefore, we chose to only keep the efficacy of the la-mAB after its administration. We tested an alternative scenario where the effects of the two interventions would combine.

## Results

### Epidemiology of RSV infection

The infection model fit the data well, capturing the difference in infection timing between children born in different seasons (Fig. 2A). The model fit diagnostics can be found in the appendix.

**Figure 2:**
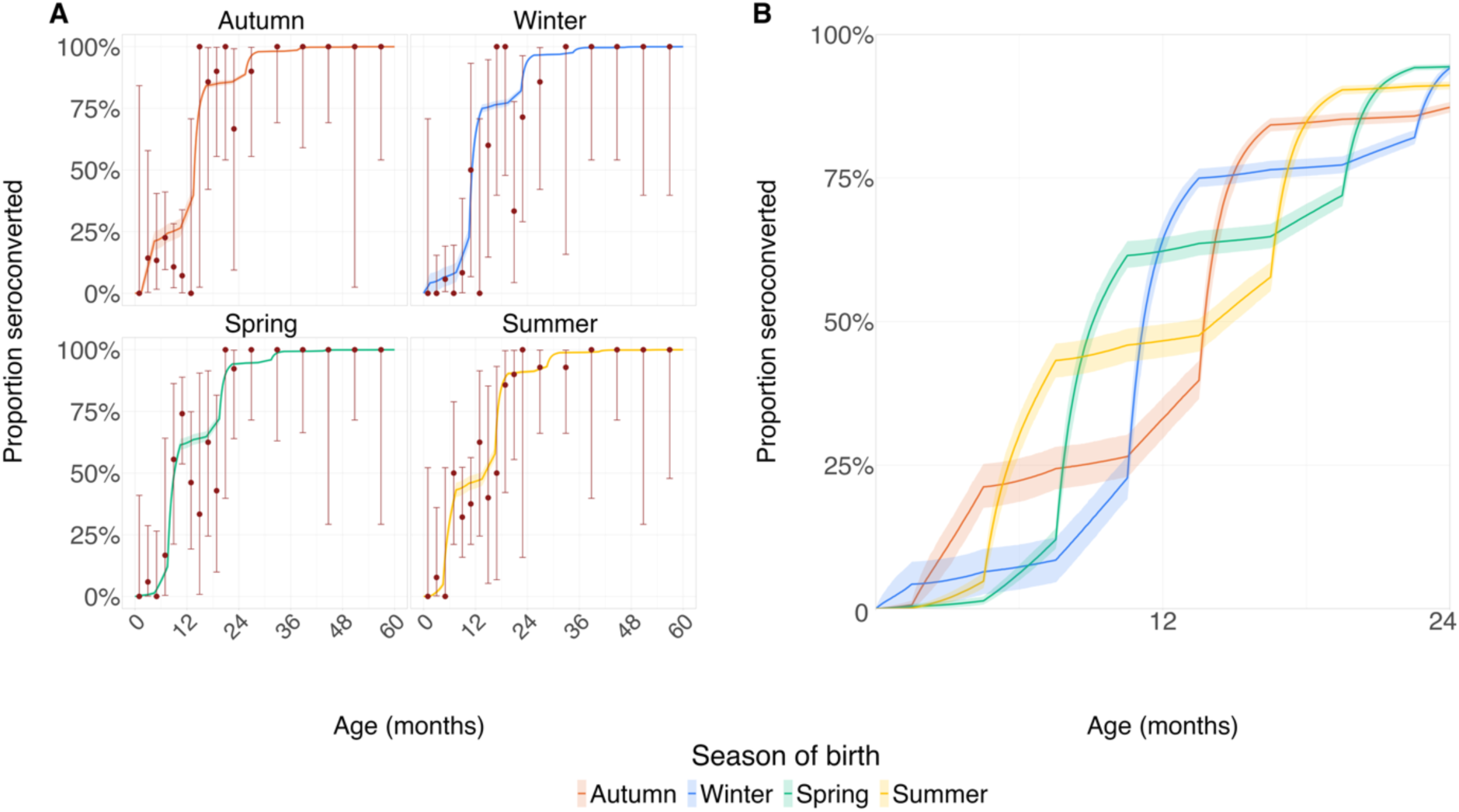
Modelled proportion of seroconverted children and seroprevalence data by season of birth The median modelled proportion of seroconverted children is shown with the corresponding 95% credible intervals (shaded areas) for ages 0-5 years (panel A) and 0-2 years (panel B). The observed seroprevalence is shown on panel A in red: dots show the point estimates and vertical bars show the 95% confidence intervals.

We estimated that 98% (95% CrI: 90-100) of children are born protected by naturally-derived maternal immunity against RSV infection. At the age of six months, 61% (95% CrI: 58 – 64) of children still had maternal protection, and 26% (95% CrI: 24 - 28) at 12 months. The probability of infection was found to vary by season, being lowest in the summer and highest in the winter with a 96% probability of being exposed at least once during winter compared to 5% in the summer (Table S2). As a result of maternal protection and seasonal risk of exposure, the proportion of infections in the first year of life also varied by birth cohort. At the age of six months, 17% (95% CrI: 14 – 20) of all children had experienced RSV infection. This was highest among autumn-born children (22%, 95% CrI: 18 – 26), while only 6% (95% CrI: 5 - 8) spring-born children had been infected at the same age. The inverse was true at 12 months of age, when 62% (95% CrI: 60 – 65) of spring-born children had seroconverted, compared to only 33% (95% CrI: 30– 37) of the autumn birth cohort. In all birth cohorts more than 86% of children had seroconverted by 2 years of age.

### Burden of RSV disease

The risk for RSV associated hospitalisation or ICU attendance if infected decreased substantially with age (Fig. 3). We estimated that in the first two months of life 8% (95% CI: 4 - 47) of RSV infected newborns would be hospitalised and 24% (95% CI: 16 – 34) of those hospitalised would be admitted to intensive care. In comparison, 3% (95% CI: 4 - 6) of 6-month-old children infected with RSV would be hospitalised and 4% (95% CI: 3 - 5) of those, admitted to ICU.

**Figure 3:**
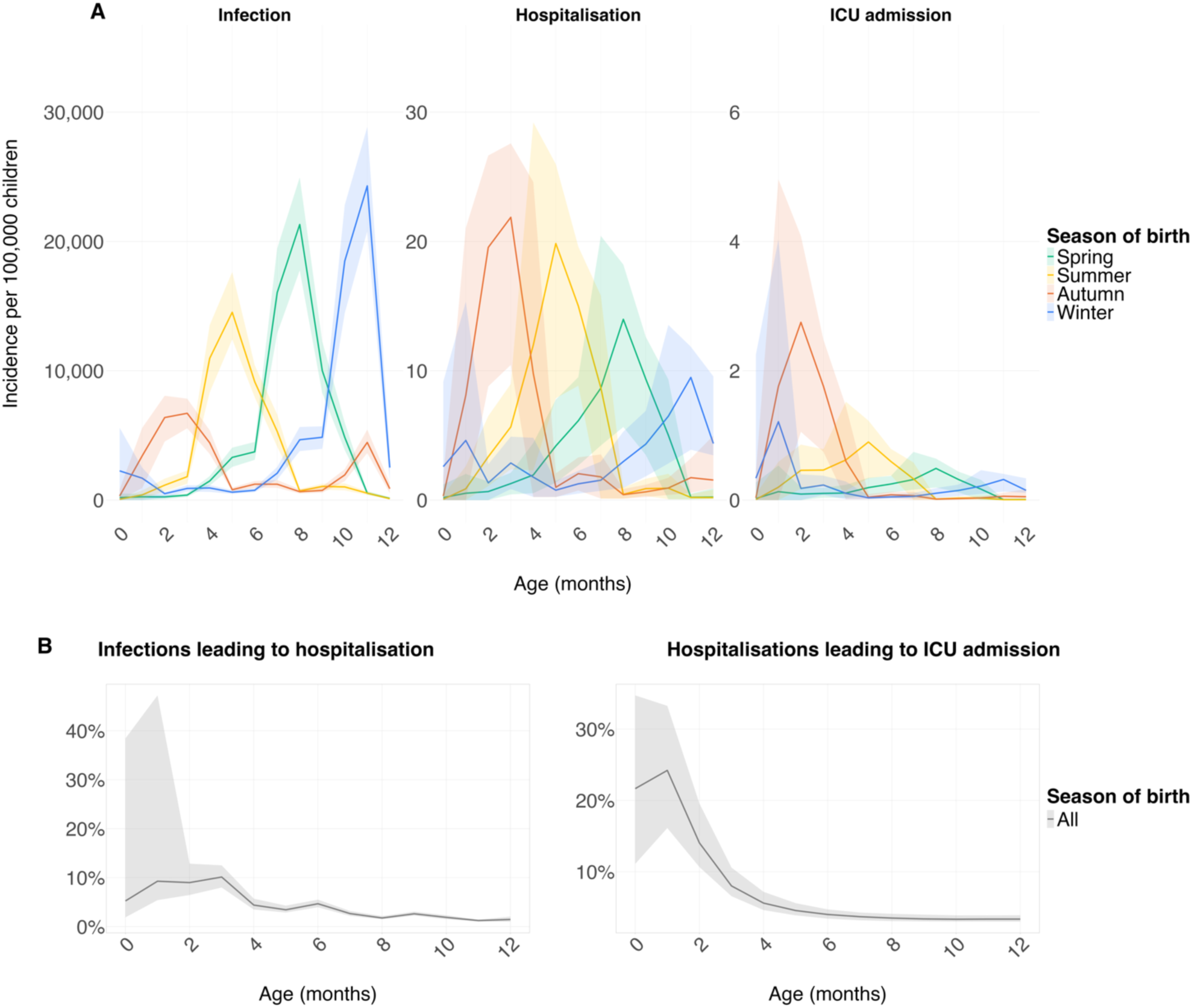
Monthly incidence of RSV outcomes in children under the age of 1 year in Germany (A) Infections, hospitalisations, and intensive care unit (ICU) admissions are shown by month of age and season of birth as central estimates and 95% confidence intervals (shaded area). (B) Disease progression by age as central estimates and 95% confidence intervals (shaded area).

The incidence of hospitalisation during the first year of life was highest in the children born in the summer (2,009/100,000, 95% CI: 1,712 – 2,283) and lowest for those born in the winter (1,300/100,000, 95% CI: 1,016 – 1,412). Hospitalisation incidence peaked at 22 (95% CI: 10 – 28) per 100,000 per month at 3 months of age in autumn-born infants and at 20 (95% CI: 8 - 26) per 100,000 per month at 5 months of age in summer-born children (Fig. 3A). The mean age at hospitalisations was estimated at 5.3 months (95% CI: 5.0 – 5.7) (Table 1). The longer before the beginning of the RSV season a cohort was born, the older their average age at hospitalisation: autumn-born children were on average hospitalised at 3.3 months (95% CI: 3.1 – 3.5) while spring-born children were, on average, hospitalised 4 months later (7.2, 95% CI: 7.0 – 7.4).

**Table 1:**
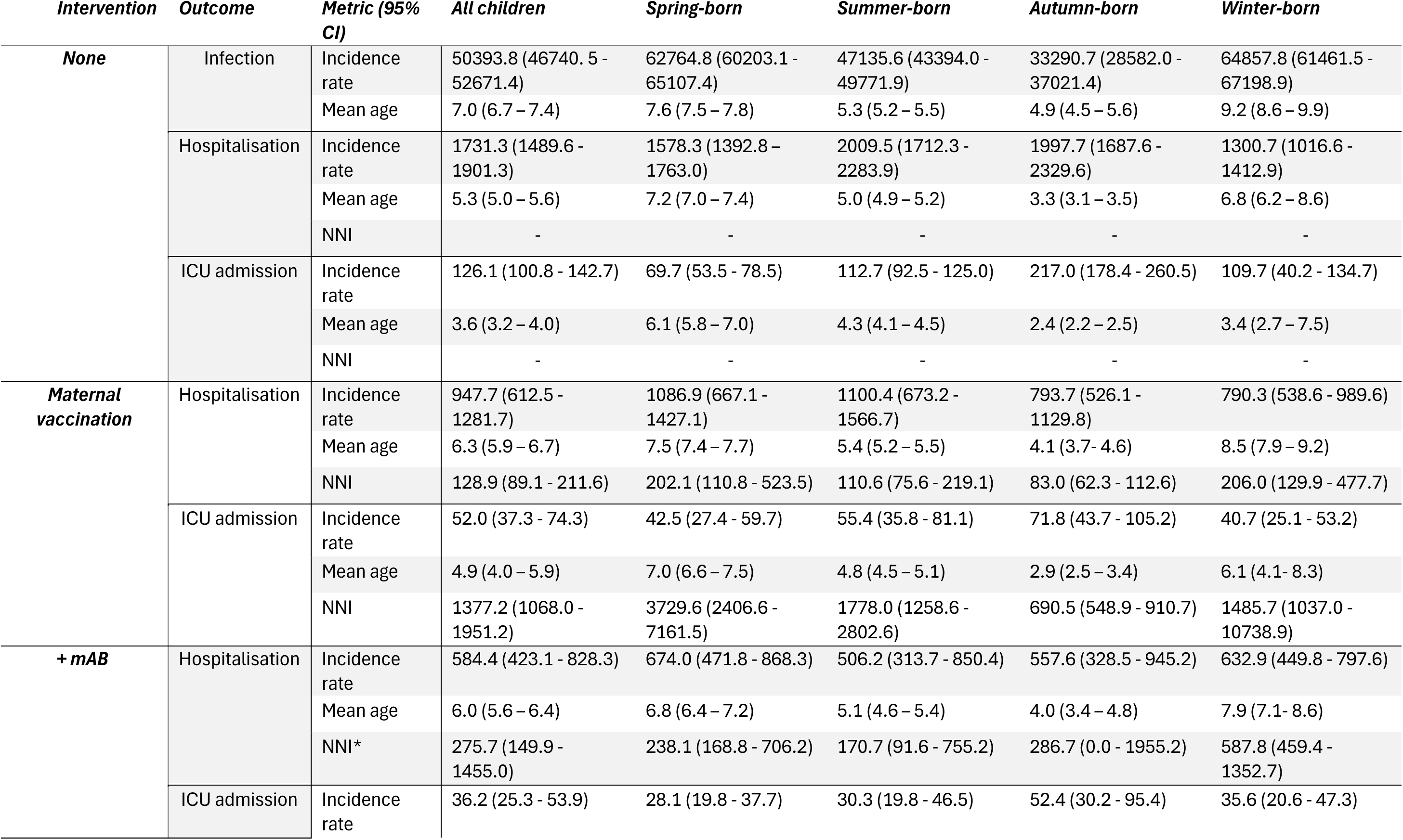
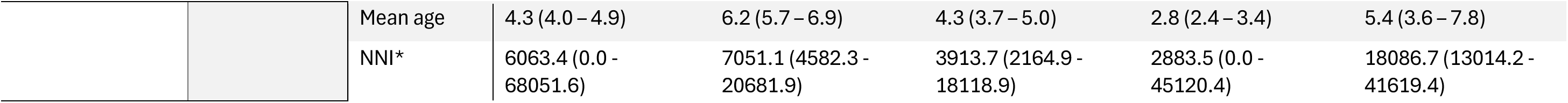
RSV-related outcomes in children <1 year old by intervention and season of birth The maternal vaccine is modelled to be given with 100% coverage. mAB administration is seasonal and universal. Rates are given per 100,000 children. The mean age is given in months. *NNI to prevent 1 hospitalisation or 1 ICU admission in the birth cohort mAB: long-acting monoclonal antibody (nirsevimab) NNI: number needed to immunise to prevent one hospitalisation / ICU admission CI: Confidence interval

The incidence of ICU admission in the first year of life was highest for autumn-born children (217/100,000, 95% CI: 178 – 260) and lowest in the spring-born cohort (70/100,000, 95% CI: 53 – 79). The peak monthly incidence in ICU admissions happened at 1 month (1.0/100,000, 95% CI: 0.0 – 1.4). At this age, peak ICU admission incidence was especially high in the autumn birth cohort at 2/100,000 (95% CI: 0 – 5). The average age at ICU admission was lower than at hospitalisation at 3.6 months (95% CI: 3.2 – 4.1) and autumn-born children were, on average, admitted earlier (2.4 months, 95% CI: 2.3 – 2.6) than those born in other seasons.

### Impact of RSV prevention

#### Impact of maternal vaccination

We estimate that MV could prevent 776 (95% CI: 473 – 1,122) infant RSV hospitalisations per 100,000 children born to vaccinated mothers and 73 (95% CI: 51 – 94) per 100,000 ICU admissions annually in Germany, a 45% (95% CI: 28 – 65) and 58% (95% CI: 43 - 72) reduction compared to no maternal immunisation (Table 1).

Autumn- and winter-born children benefit most from MV. We estimate that MV can prevent 145 (95% CI: 110 - 182) per 100,000 or 66.53% (95% CI: 53.32 - 77.43) of RSV associated ICU admissions among autumn-born children and 67 (95% CI: 9 - 96) per 100,000 or 61% (95% CI: 25 - 73) among winter-born children. While benefitting less, at 43 (95% CI: 27 - 60) and 55 (95% CI: 36 - 81) per 100,000 the incidence of ICU admission in the first year of life among children born to vaccinated mothers in spring and summer was substantially lower than in children born to vaccinated mothers in autumn and comparable to winter-born children (Table 1).

Similarly, the autumn birth cohort also had the greatest reduction in hospitalisations following MV, with 793 (95% CI: 526 - 1130) per 100,000 or 60% (95% CI: 45 - 74) of cases prevented. The smallest impact was seen among spring-born children, where only 32% (95% CI: 12 - 57) of hospitalisations were prevented by the MV.

#### Impact of added la-mAB

If given to infants as they enter their first RSV season, la-mAB would prevent an additional 360 (95% CI: 52 - 638) hospitalisations and 16 (95% CI: 0 – 35) ICU admissions per 100,000 children born to RSV vaccinated mothers; a 36% (95% CI: 8 - 54) and 30% (95% CI: 0 - 51) increase in impact over what would be prevented by MV alone.

Administration of la-mAB to the summer-born cohort would result in the biggest relative reduction (52% reduction in hospitalisations (95% CI: 12 - 72), 46% reduction in ICU admissions (95% CI: 10 - 63)) (Fig. 4) compared to MV alone, albeit corresponding to only a modest absolute reduction of 575 (95% CI: 94 – 1,017) hospitalisations and 25 (95% CI: 4 – 45) ICU admissions prevented per 100,000 additionally immunised children born in this season. The impact on ICU admissions was smaller than the impact on hospitalisations as this birth cohort would receive the la-mAB at 3 months, after the peak in ICU admissions. In spring-born infants, the la-mAB would reduce hospitalisations by an additional 39% (95% CI: 19 - 47) and ICU admissions by an additional 33% (95% CI: 16 - 41), the decrease being 420 (95% CI: 142 – 593) per 100,000 hospitalisations and 14 (95% CI: 5 – 22) per 100,000 ICU admissions.

**Figure 4:**
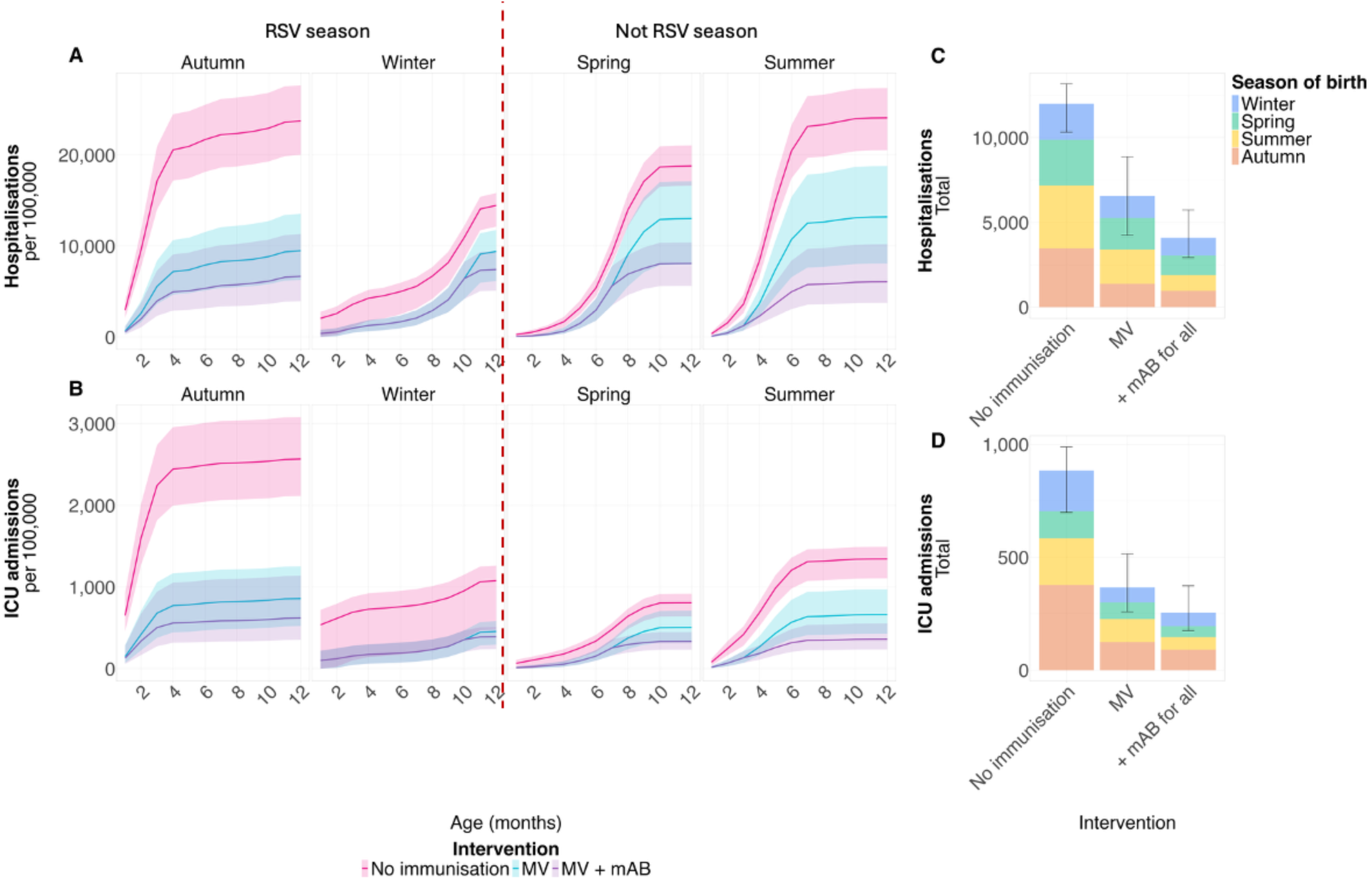
Cumulative incidence of RSV outcomes in children under the age of 1 year Panels A and B respectively show cumulative hospitalisation and intensive care unit (ICU) admission incidences per 100,000 by month of age and season of birth as central estimates and 95% confidence intervals (shaded area). Panels C and D respectively show the total number of hospitalisations and ICU admissions by intervention and season of birth. The vertical lines represent the 95% confidence interval. The long-acting monoclonal antibody (mAB) is given to children right before the start of their first RSV season.

#### Efficiency of immunisation

An estimated 129 (95% CI: 90 - 212) mothers would need to be vaccinated to prevent one infant RSV hospitalisation and 1,378 (95% CI: 1,069 – 1,952) to prevent one ICU admission. The NNI to prevent one ICU admission in the spring and summer birth cohorts was 3,730 (95% CI: 2,407 – 7,162) and 1,779 (1,259 – 2,803) (Table 1).

In addition to MV, 171 (95% CI: 92 - 756) la-mAB doses would need to be administered pre-season to infants born in the summer and 239 (95% CI: 169 - 706) to spring-born children to prevent one additional hospitalisation. One ICU admission would be avoided in those birth cohorts if 3,914 (95% CI: 2,165 – 18,119) and 7,052 (95% CI: 4,583 – 20,682) children respectively received an additional la-mAB dose.

### Sensitivity analysis

We found little evidence for birth season-specific protection by naturally-derived maternal immunity. Changing the structure of the infection model to simulate maternal immunity waning following an Erlang-3 distribution did also not change the estimated parameter values in the infection model. Assuming an exponential waning rate (Erlang-1) shortened the estimated duration of immunity slightly to 8 months. The corresponding values can be found in the supplementary material.

Under the assumption that the effect of the MV and of the la-mAb combine instead of the la-mAB replacing the remainders of the protection offered by MV, the number of hospitalisations would decrease by 22% (95% CI: 10 - 40) while the number of ICU admissions would increase by 29% (95% CI: 16 - 45) (Table S7). Children born in the autumn, who would receive the la-mAB early in life when protection from the MV is still high, would see the biggest difference in estimated impact compared to our base case assumptions.

## Discussion

Children born in the summer and the autumn, i.e., shortly before the start of the RSV season, have the highest disease burden in the first year of life with about 2,000 hospitalisations and over 100 ICU admissions per 100,000 children. This reflects the combination of waning of maternally derived immunity, seasonal patterns in exposure risk, and decreasing risk of severe disease for infections later in infancy.^33–36^ MV would substantially reduce the disease burden, preventing over 500 ICU admissions per 100,000 children born to vaccinated mothers, almost 60%. The benefit is disproportional in children born in the autumn and the winter, with over 60% reduction in ICU admissions, but MV would bring the risk of severe disease to comparable levels across all birth cohorts. Additional administration of the la-mAB would protect infants even further, particularly those born in the spring and the summer, but the reduction is incidence is relatively small and the NNIs much higher than in the maternal programme.

An important result of our study is the difference between hospitalisation and ICU admission risk within birth seasons. ICU admissions peak earlier in life than hospitalisations, which is reflected by the more important relative reduction in ICU cases after MV. This is especially striking for children born in the winter, for whom MV would have little impact on hospitalisation risk, but divide the risk of ICU admission by two. Conversely, as the la-mAB would be given later in life, it would primarily reduce hospitalisation incidence, regardless of birth season.

Our findings highlight that additional la-mAB administration would not prevent substantial disease burden, especially if not timed strategically. Further, implementing a seasonal schedule could prove difficult given the challenge of predicting the start of the RSV season – particularly in the post-COVID-19 era. RSV seasonality has shifted in several countries, including Germany, as the peak in RSV cases was shown to happen earlier in the two seasons following the pandemic than in the pre-pandemic period. ^37–39^ As our results are based on single season data collected before the COVID-19 outbreak, they would need to be revised if these trends were to continue, with children born in the spring possibly benefiting from the interventions more than those born in the autumn under such conditions.

Our birth season specific RSV outcome estimates match population-based observations from the few studies separating outcomes for children under the age of one year into monthly groups^40,41^. These studies used observations from cohort studies and hospitalisation records originating from Croatia and the USA respectively to examine the role of birth of month in age at first RSV infection and risk of severe disease. In accordance with our study, they show that children born in the autumn and at the beginning of winter experience their first infection at an earlier age than other infants and have a higher risk of hospitalisation.

Our findings are not based on Germany data only and are therefore generalisable to the other countries where both immunisation products are available. For example, the United Kingdom expanded its strategy in the 2025/2026 season to recommend nirsevimab administration to premature and other high-risk infants born to vaccinated mothers.^42^ Our results suggest that extending this recommendation to other infants would provide modest additional benefit, even if focusing on children born just before the start of the RSV season. In addition, our study shows that, in countries where MV is the preferred immunisation route, la-mAB could be used to protect children if coverage rates are low, e.g. in the USA.^43,44^ Indeed, the small benefit shown here assumes a complete vaccination coverage, so a higher impact is likely in low MV uptake settings.

Some limitations of our study are worth noting. We calibrated our model using data originating from outside Germany. However, the epidemiology of RSV in the Netherlands is likely comparable to Germany given their close geographical proximity and therefore their climatic similarity.^45,46^ We use age stratified estimates for hospitalisation and ICU admission incidence from Mahmud *et al*^21^ to estimate how the burden of disease is divided among age groups in the first year of life. While these estimates are based on data from low- and middle-income countries, the author report no evidence for their estimates to vary with income group. In addition, while granular data is not available for Germany, the age distribution of hospitalised cases and ICU admissions in our model is consistent with published in German reports.^4^ Furthermore, our model does not account for pre-term births, contact with older siblings in the household, or comorbidities, all of which are known risk factors for RSV hospitalisations.^9,33,47^ As a result, we cannot disentangle the benefit of MV or additional administration of la-mAB to premature infants and other high-risk groups, but instead provide estimates for a population average. While MV was shown to also provide adequate protection to pre-term infants, children born within two weeks after maternal immunisation or with other underlying risk factors for severe outcomes would likely benefit disproportionally from la-mAB. ^42,44,48^ Thirdly, we modelled a birth cohort in which all mothers would be immunised. As coverage for maternal immunisation in Germany and other European countries are typically well below 80%^49,50^, any estimates of number of cases prevented are to be understood as best case scenario estimates and not be mistaken for real life predictions. However, we mostly report estimates for the reduction in incidence per 100,000 children born to immunised mothers or relative risk. Given that only minor indirect effects of the programme are expected, these are independent of population level coverage assumptions. Finally, our 2023 baseline disease burden reflects the previous standard of care (administration of palivizumab to high-risk infants), not a no-intervention scenario, making this a comparison of immunisation strategy alternatives rather than comparison to no intervention.

## Conclusion

Our study highlights the strong heterogeneity in the risk of severe RSV disease across seasonal birth cohorts. MV greatly reduces and homogenises the disease incidence across birth seasons. Additional administration of the la-mAB before the start of the season, particularly to those born in spring and summer and thus with little residual protection from maternal vaccine as they enter their first RSV season, further lowers the risk for severe RSV. However, the benefit is modest, and this strategy represents a less efficient use of immunisation if compared to the MV programme.

## Data Availability

All the data used for this analysis is publicly available. Seroprevalence data is available from https://github.com/Stijn-A/RSV_serology. German hospitalisation numbers are available from https://datenbrowser.inek.org/. Disease progression numbers are available from Mahmud et al (https://doi.org/10.1016/S2352-4642(25)00349-9). Immunisation efficacy estimates were Monoi et al (https://github.com/ayakamon/BR-RSV-MV and https://github.com/ayakamon/RSV_SEA_IMMUN).

https://github.com/jmayer-cgh/RSV_MV_mAB_catchup

## Contributors

JM conceptualised the study with input from SF. JM led data analysis with input from SF, BK, and MD. JM wrote the codes for analysis with input from FK, AM, and KvZ. JM and SF led the data interpretation. JM wrote the first draft of the manuscript with input from SF, AM, BK, and MD. All authors contributed to data interpretation and critically revised the manuscript. All authors read and approved the final version of the manuscript. JM and SF had full access to and verified the underlying data of the study. All authors had final responsibility for the decision to submit for publication.

## Data sharing statement

All the data used for this analysis is publicly available. Seroprevalence data is available from https://github.com/Stijn-A/RSV_serology. German hospitalisation numbers are available from https://datenbrowser.inek.org/. Disease progression numbers are available from Mahmud *et al*^21^. Immunisation efficacy estimates were derived from Kampmann *et al*^16^ and Hammit *et al*^25^.

The model code can be found at https://github.com/jmayer-cgh/RSV_MV_mAB_catchup.

## Declaration of interests

The authors declare no conflicts of interest related to the present study. SF is a member of the German NITAG (STIKO).

