## Supplementary material for "Effectiveness and efficiency of pre-season administration of long-acting monoclonal antibodies for infants born to RSV vaccinated mothers: a modelling study"

**Model structure**

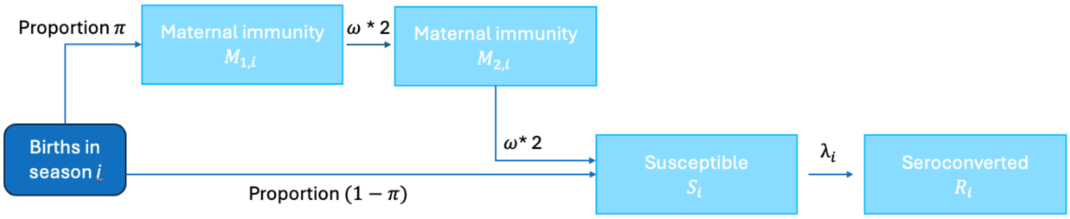

**Figure S1**: **Structure of the catalytic model**

In each season of the year *i,* a proportion π, of children is born with maternal immunity against RSV which wanes with a rate ω according to an Erlang-2 distribution ($M_1$ and $M_2$ compartments). Susceptible children (S) experience a seasonal force of infection $\lambda_j$ which leads them to seroconvert (R).

**Fitting procedure**

The catalytic model is fitted to seroprevalence data using the monty package.[1] We estimate six parameters: the four seasonal components of the force of infection, the proportion of children born with maternal immunity π, and the waning rate of maternal immunity ω. The assumptions on the prior distributions and initial values are presented in table S1. We used non-informative priors with arbitrary initial values for the four components of the FOI and π. We assumed that the waning rate of maternal immunity follows a Normal distribution centred around 181 days and with 95% confidence interval [129 - 307] based on the literature.[2-5]

| Parameter | Prior distribution | Initial value |
|---|---|---|
| Summer component of the FOI ($\lambda_{summer}$) | Uniform (0, 0.1) | 0.02002 |
| Autumn component of the FOI ($\lambda_{autumn}$) | Uniform (0, 0.1) | 2.00E-05 |
| Winter component of the FOI ($\lambda_{winter}$) | Uniform (0, 0.1) | 3.00E-05 |
| Spring component of the FOI ($\lambda_{spring}$) | Uniform (0, 0.1) | 1.00E-05 |
| ω | Normal(1/181, 0.0011546) | 0.004 |
| $\pi$ | Uniform (0, 1) | 0.5 |

**Table S1: Assumptions on the prior distributions and initial values of the parameters estimated by the catalytic model.**

The force of infection (FOI) in season *i* is defined as $FOI_i = \lambda_{summer} + \lambda_i$ where $\lambda_i$=0 if i = summer.

ω: waning rate of naturally-derived maternal immunity

$\pi$: proportion of children born with naturally-derived maternal immunity

We used two steps for the fitting routine. First, we fitted the model using a naïve variance-covariance matrix C= diag(0.0004, 0.0004, 0.0004, 0.0004, 0.0004, 0.0004). We used an adaptive MCMC with 80,000 steps and 2 chains. We then used the covariance of the draws from this sample to build a new variance-covariance matrix that we used to fit the model again. We used an adaptive MCMC algorithm with 80,000 steps and 6 chains.

We extracted the estimated distribution of the six parameters from the second fit and used to simulate the proportion of seroconverted children by season of birth over time.

After a burn-in period of 3,000 steps, the chains were mixing well as shown in Figure S1. We used the effective sample size and R hat to assess convergence. The values are presented in table S2. Finally, we examined the uncertainty around the parameters estimates as shown in Figure S2.

| Variable | Median | 2.5% quantile | 97.5% quantile | R hat | ESS bulk | ESS tail |
|---|---|---|---|---|---|---|
| Summer component of the FOI ($\lambda_{summer}$) | 5.54E-04 | 2.86E-05 | 2.27E-03 | 1.00 | 3.02E+03 | 1.57E+04 |
| Autumn component of the FOI ($\lambda_{autumn}$) | 2.52E-03 | 1.84E-04 | 8.32E-03 | 1.02 | 2.38E+02 | 2.69E+03 |
| Winter component of the FOI ($\lambda_{winter}$) | 3.47E-02 | 8.76E-03 | 9.38E-02 | 1.01 | 6.18E+02 | 8.21E+02 |
| Spring component of the FOI ($\lambda_{spring}$) | 1.03E-03 | 4.30E-05 | 5.23E-03 | 1.00 | 3.08E+03 | 1.53E+04 |
| ω | 3.28E-03 | 2.75E-03 | 5.56E-03 | 1.01 | 5.36E+02 | 4.88E+02 |
| $\pi$ | 9.76E-01 | 9.04E-01 | 9.99E-01 | 1.00 | 2.76E+03 | 1.85E+03 |

**Table S2**: **Values of the four parameters estimated by the catalytic model**
The force of infection (FOI) in season $i$ is defined as $FOI_{\mathrm{i}} = \lambda_{summer} + \lambda_i$ where $\lambda_i$=0 if i = summer.
ω: waning rate of naturally-derived maternal immunity
$\pi$: proportion of children born with naturally-derived maternal immunity
ESS: essential sample size

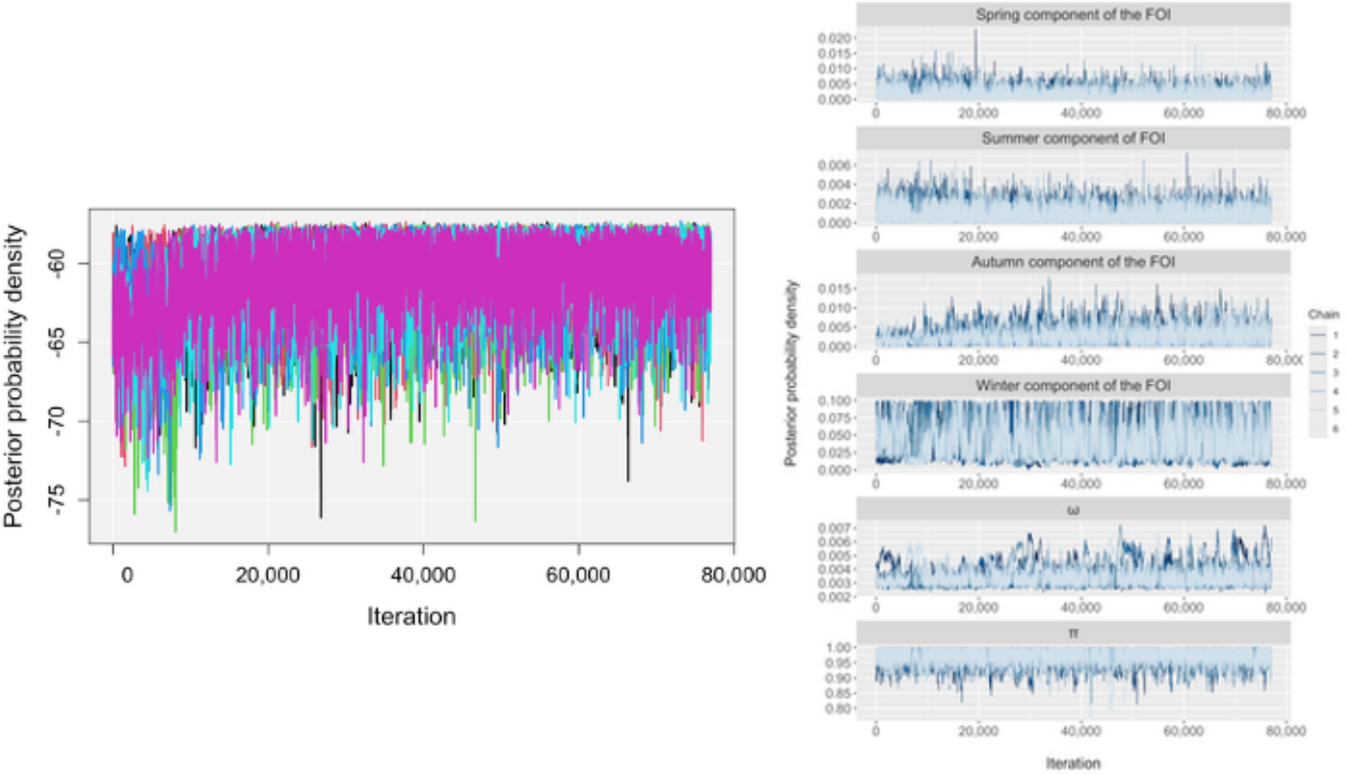

**Figure S1**: **Trace plots of the catalytic model**
The catalytic model was fitted using an adaptive MCMC with 6 chains and 80,000 iterations. The burn-in period was set to 3,000 steps. We checked the overall mixing (right panel) as well as each of the estimated parameters (left panel).
The force of infection (FOI) in season $i$ is defined as $FOI_i = \lambda_{summer} + \lambda_i$ where $\lambda_i$=0 if i = summer.
ω: waning rate of naturally-derived maternal immunity
$\pi$: proportion of children born with naturally-derived maternal immunity

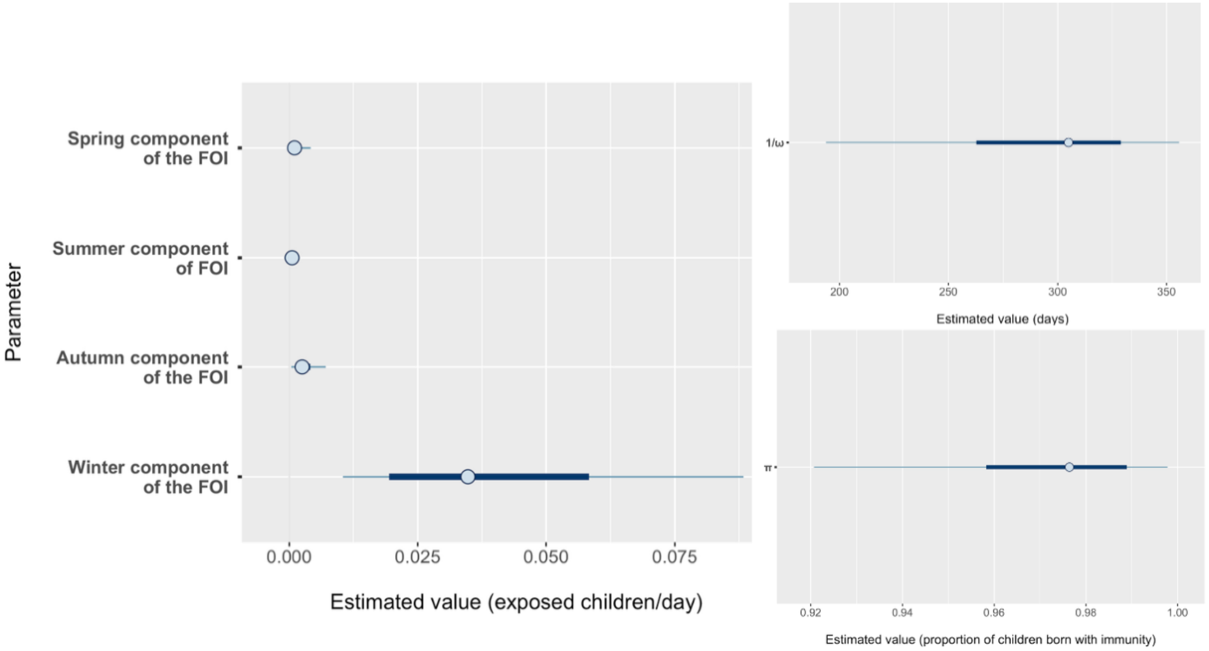

**Figure S2**: **Posterior uncertainty intervals of the parameters estimated by the catalytic model**
Points show the posterior median, the thick lines show the 50% intervals, and the thin lines show the 90% intervals.
The force of infection (FOI) in season $i$ is defined as $FOI_{\mathrm{i}} = \lambda_{summer} + \lambda_i$ where $\lambda_i$=0 if i = summer.
ω: waning rate of naturally-derived maternal immunity
$\pi$: proportion of children born with naturally-derived maternal immunity

### Sensitivity analyses

#### Exponential waning of maternal immunity

To test the sensitivity of the model to the assumption of maternal immunity waning following a second order Erlang distribution, we tried an alternative formulation with only one M compartment, modelling an exponential waning of immunity. This resulted in a shorter protection window (231 days, 95% CrI: 146 – 342) than in the baseline model, although the 95% CrI overlapped. The convergence of the model was slightly worse, as indicated by the higher R hat and ESS values (Table S3).

| Variable | Median | 2.5% quantile | 97.5% quantile | R hat | ESS bulk | ESS tail |
|---|---|---|---|---|---|---|
| Summer component of the FOI ($\lambda_{summer}$) | 3.52E-04 | 1.65E-05 | 1.43E-03 | 1.01 | 2.62E+03 | 2.78E+03 |
| Autumn component of the FOI ($\lambda_{autumn}$) | 3.44E-03 | 5.57E-04 | 1.28E-02 | 1.04 | 1.22E+02 | 1.06E+02 |
| Winter component of the FOI ($\lambda_{winter}$) | 1.21E-02 | 2.07E-03 | 4.12E-02 | 1.03 | 1.62E+02 | 1.19E+02 |
| Spring component of the FOI ($\lambda_{spring}$) | 5.17E-04 | 2.00E-05 | 2.56E-03 | 1.01 | 3.44E+03 | 2.58E+03 |
| ω | 4.32-03 | 2.92E-03 | 6.84E-03 | 1.02 | 3.60E+02 | 3.58E+02 |
| $\pi$ | 9.83E-01 | 8.68E-01 | 9.99E-01 | 1.01 | 8.68E+03 | 3.42E+03 |

**Table S3: Values of the four parameters estimated by the catalytic model under the assumption of exponential maternal antibody decay**

The force of infection (FOI) in season $i$ is defined as $FOI_i = \lambda_{summer} + \lambda_i$ where $\lambda_i$=0 if i = summer.
ω: waning rate of naturally-derived maternal immunity
$\pi$: proportion of children born with naturally-derived maternal immunity
ESS: essential sample size

### Additional M sub-compartment

We tried adding a third M sub-compartment M3 to have maternal immunity wane following an Erlang-3 distribution. As shown on figure S3 and table S4, this did not improve the model fit or change the estimates parameter values, so we chose to keep an Erlang-2 distribution for the lower computational cost.

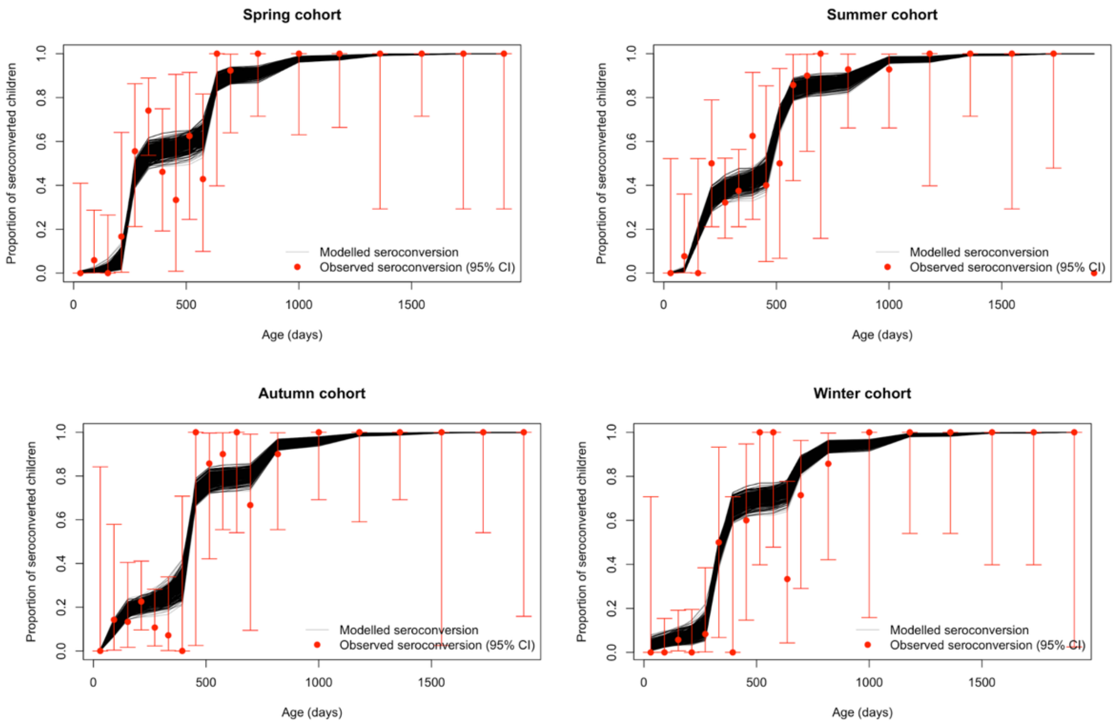

**Figure S3: Model fit to the seroprevalence data by season of birth using three M sub-compartments**
The catalytic model was fitted using an adaptive MCMC with 6 chains and 80,000 iterations. The seroprevalence data was divided into four birth cohorts to match the four yearly seasons. The modelled proportion of seroconverted children is shown for each iteration, as well the observed seroprevalence and the corresponding 95% confidence interval (CI).

| Variable | Median | 2.5% quantile | 97.5% quantile | R hat | ESS bulk | ESS tail |
|---|---|---|---|---|---|---|
| Summer component of the FOI ($\lambda_{summer}$) | 6.46E-04 | 4.71E-05 | 1.83E-03 | 1.00 | 5.14E+03 | 7.47E+03 |
| Autumn component of the FOI ($\lambda_{autumn}$) | 2.85E-03 | 4.26E-04 | 7.53E-03 | 1.00 | 4.01E+03 | 5.62E+03 |
| Winter component of the FOI ($\lambda_{winter}$) | 3.58E-02 | 9.99E-03 | 9.02E-02 | 1.01 | 5.23E+02 | 7.74E+02 |
| Spring component of the FOI ($\lambda_{spring}$) | 1.29E-03 | 1.05E-04 | 6.13E-03 | 1.00 | 4.71E+03 | 1.23E+04 |
| ω | 3.25E-03 | 2.78E-03 | 5.36E-03 | 1.01 | 5.45E+02 | 4.69E+02 |
| $\pi$ | 9.78E-01 | 9.24E-01 | 9.98E-01 | 1.00 | 4.35E+03 | 2.40E+03 |

**Table S4**: **Values of the four parameters estimated by the catalytic model with 3 M subcompartments**
**The force of infection (FOI) in season *i* is defined as $FOI_i = \lambda_{summer} + \lambda_i$ where $\lambda_i$=0 if i = summer.**
ω: waning rate of naturally-derived maternal immunity
$\pi$: proportion of children born with naturally-derived maternal immunity

### Non-informative prior for the waning of maternal immunity

Using a flat, non-informative prior for the waning rate of maternal immunity ω when fitting the model resulted in similar parameter value estimates but worse performance of the MCMC (table S5). We used ω ~ Uniform (0.0001, 0.1) with initial value 0.004. All other priors were the same as shown in Table S1.

| Variable | Median | 2.5% quantile | 97.5% quantile | R hat | ESS bulk | ESS tail |
|---|---|---|---|---|---|---|
| Summer component of the FOI ($\lambda_{summer}$) | 4.93E-04 | 3.56E-05 | 2.42E-03 | 1.00 | 2.05E+03 | 6.29E+03 |
| Autumn component of the FOI ($\lambda_{autumn}$) | 2.78E-03 | 2.28E-04 | 8.80E-03 | 1.02 | 4.28E+02 | 1.63E+03 |
| Winter component of the FOI ($\lambda_{winter}$) | 2.11E-02 | 8.46E-03 | 8.66E-02 | 1.23 | 1.82E+01 | 1.35E+02 |
| Spring component of the FOI ($\lambda_{spring}$) | 1.08E-03 | 4.55E-05 | 5.03E-03 | 1.01 | 2.43E+03 | 1.61E+04 |
| ω | 3.47E-03 | 2.75E-03 | 5.00E-03 | 1.15 | 2.63E+01 | 1.87E+02 |
| $\pi$ | 9.70E-01 | 8.89E-01 | 9.99E-01 | 1.01 | 6.57E+02 | 1.63E+03 |

**Table S5**: **Values of the four parameters estimated by the catalytic model with 2 M sub-compartments using a Uniform prior for ω**

The force of infection (FOI) in season *i* is defined as $FOI_i = \lambda_{summer} + \lambda_i$ where $\lambda_i$=0 if i = summer.

ω: waning rate of naturally-derived maternal immunity

$\pi$: proportion of children born with naturally-derived maternal immunity

### Season-dependent maternal protection

Given the strong seasonality of RSV, we tested an alternative hypothesis where the proportion of children born with naturally-derived maternal immunity is dependent on the season of birth. This assumes that only mothers exposed to RSV shortly before giving birth would pass on AB to their children.[6] The resulting estimates suggested that fewer children born in the spring would be protected at birth compared to the other seasons. However, the model struggled to converge for this parameter (table S6).

| Variable | Median | 2.5% quantile | 97.5% quantile | R hat | ESS bulk | ESS tail |
|---|---|---|---|---|---|---|
| Summer component of the FOI ($\lambda_{summer}$) | 4.63E-04 | 1.85E-05 | 2.25E-03 | 1.03 | 1.37E+02 | 5.85E+02 |
| Autumn component of the FOI ($\lambda_{autumn}$) | 2.52E-03 | 1.74E-04 | 8.31E-03 | 1.03 | 1.73E+02 | 5.76E+02 |
| Winter component of the FOI ($\lambda_{winter}$) | 4.84E-02 | 6.46E-03 | 9.86E-02 | 1.22 | 1.95E+01 | 1.49E+02 |
| Spring component of the FOI ($\lambda_{spring}$) | 1.42E-03 | 5.95E-05 | 8.00E-03 | 1.02 | 1.74E+02 | 1.87E+03 |
| ω | 3.01E-03 | 2.56E-03 | 5.87E-03 | 1.08 | 4.66E+01 | 1.81E+02 |
| $\pi_{spring}$ | 8.55E-01 | 3.05E-01 | 9.90E-01 | 1.20 | 2.09E+01 | 1.74E+02 |
| $\pi_{summer}$ | 9.43E-01 | 7.42E-01 | 9.97E-01 | 1.07 | 5.67E+01 | 2.22E+02 |
| $\pi_{autumn}$ | 9.69E-01 | 8.19E-01 | 9.99E-01 | 1.01 | 5.57E+02 | 1.99E+02 |
| $\pi_{winter}$ | 9.77E-01 | 8.05E-01 | 9.99E-01 | 1.03 | 2.10E+02 | 1.75E+02 |

**Table S6**: **Values of the four parameters estimated by the catalytic model with a season-specific protection by maternal immunity**

The force of infection (FOI) in season *i* is defined as $FOI_i = \lambda_{summer} + \lambda_i$ where $\lambda_i$=0 if i = summer.

ω: waning rate of naturally-derived maternal immunity

$\pi_i$: proportion of children born with naturally-derived maternal immunity in season *i*

### Cumulative effect of immunisation protection

The assumption of a cumulative effect of the MV and the mAB produced lower numbers of RSV-related outcomes. As expected, the decrease in the number of outcomes was highest for the cohort who would receive the intervention early in life when the VE is still high. The difference was negligible for children born in the winter, modelled to receive the la-mAB at 11 months.

| Season of birth | Decrease in hospitalisations (95% CI) | Decrease in ICU admissions (95% CI) |
|---|---|---|
| Spring | 5.24% (0.83 - 19.08) | 4.27% (0.71 - 15.23) |
| Summer | 31.44% (12.43 - 52.33) | 25.29% (9.14 - 44.36) |
| Autumn | 55.4% (38.29 - 70.35) | 63.15% (47.71 - 74.88) |
| Winter | 1.00% (0.09 - 5.79) | 0.60% (0.06 - 2.94) |
| All | 22.09% (10.36 - 39.77) | 28.66% (16.28 - 45.18) |

**Table S7**: **Reduction in RSV-related outcomes under the cumulative effect assumption compared to baseline**

The baseline model assumes that children are protected by the monoclonal antibody only after its administration. The alternative model assumes that the protection of the two immunisation products combines.

CI: confidence interval

ICU: intensive care unit

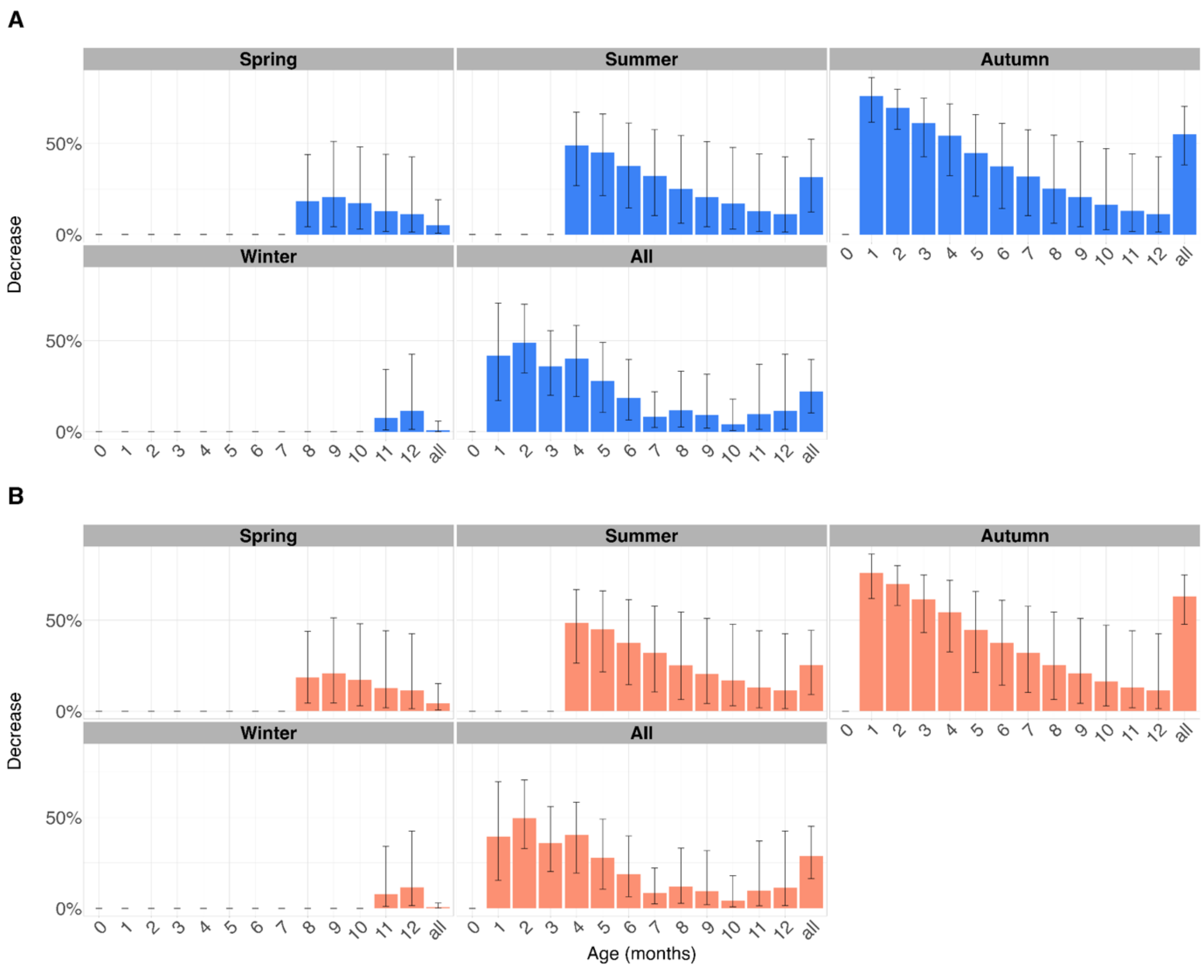

**Figure S4**: **Reduction in RSV-related hospitalisations (A) and ICU admissions (B) under the cumulative effect assumption compared to baseline**

The baseline model assumes that children are protected by the monoclonal antibody (la-mAB) only after its administration. The alternative model assumes that the protection of the two immunisation products combines. The age at administration of the mAB is birth season-specific: 1 month, 4 months, 8 months, and 11 months for autumn, summer, spring, and winter born children respectively.

The height of the bars shows points estimates, vertical lines show 95% confidence intervals.

ICU: Intensive care unit
